# Prediction of arrhythmogenesis in non-ischemic Cardiac Resynchronization Therapy patients

**DOI:** 10.64898/2025.12.15.25342270

**Authors:** Alessandra Corda, Massimiliano Maines, Stefano Pagani, Domenico Catanzariti, Maurizio Del Greco, Christian Vergara

## Abstract

**Aims:** We aim to develop a patient-specific computational model to predict the risk of Ventricular Tachycardia (VT) in patients with Biventricular Cardiac Resynchronization Therapy (BiV-CRT) device. Patients are indeed at risk of developing arrhythmias due to BiV-CRT pacing, a known potential complication that puts the cardiologist on guard against its prevention.

**Materials and Methods:** We consider three non-ischemic fibrotic patients. Patient-specific left ventricle geometries and fibrosis regions are extracted from Cine-MRI and LGE-MRI. The electrophysiology model, based on the monodomain equation and on the Ten Tusscher-Panfilov (TTP06) ionic current model, is personalized using pre-operative Electro-Anatomical Mappings System data. The TTP06 parameters are adapted to reflect the altered electrical properties of the fibrotic tissue. To test inducibility, we use an 𝕊1−𝕊2 stimulation protocol: 𝕊1 simulates the clinical BiV-CRT pacing with patient-specific VV-delay, followed by a 𝕊2 ectopic impulse. This procedure is repeated for ten ectopic sites. The arrhytmogenic risk is quantified by the number of ectopic sites that successfully generates a reentry loop.

**Conclusion:** The model’s prediction of VT risk is consistent with the long-term clinical follow-up for all the patients. Arrhythmic patients show a higher number of ectopic sites from which a reentry loop is generated compared to the non-arrhythmic patient. This study provides a first, preliminary attempt towards the use of computational tools in assessing the vulnerability of the arrhythmic substrate during BiV-CRT pacing in non-ischemic patients. In future, such tools could serve as a powerful non-invasive diagnostic metric to inform clinicians about possible therapies to associate to BiV-CRT.

## 1 Introduction

*Biventricular Cardiac Resynchronization Therapy* (BiV-CRT) represents a clinical standard of care for patients suffering from ventricular dyssynchrony, a condition that impairs the synchronism of the cardiac contraction causing heart failure. By restoring the regular ventricular motion, BiV-CRT aims to improve the systolic function and cardiac output [1]. Despite significant advances in the pacing field, such as Conduction System Pacing [2], traditional BiV-CRT remains one of the preferred first-line treatment for cardiac resynchronization [3].

However, BiV-CRT carries important risks that can lead to adverse outcomes, such as arrhythmogenesis [4, 5]. Indeed, BiV-CRT may introduce electrical instabilities due to its intrinsic non-physiological activation. This induces a reversal of the physiological endo-to-epi depolarization direction to an epi-to-endo direction, which is hypothesized to promote heterogeneities during repolarization. This could prolong the QT interval and consequently result in malignant arrhythmias [6, 7, 8]. Such a risk is in BiV-CRT patients furtherly increases by the presence of ischemic or non-ischemic fibrosis [8, 9, 10].

Given these risks, it would be clinically important to develop reliable methods for identifying which patients included in the CRT program could face an elevated arrhythmic propensity after the implantation. In this way, clinicians could assess wheter it would be advisable to implant a *CRT Defibrillator* (CRT-D) device [3].

Computational cardiac models are able to produce the action potential along the whole heartbeat in virtual, clinically-relevant, scenarios of a patient [11, 12, 13]. Specifically, over the last decade, computational research focused on patient-specific models, particularly for individuals with myocardial fibrosis [14]. This allowed to develop several computational studies with the aim of analysing or improving BiV-CRT: evaluating acute hemodynamic changes associated with the stimulation [15]; identifying non-responders through detailed electro-mechanical simulations [16, 17]; optimizing electrode placement and VV-delay [18, 19, 20, 21, 22]; evaluating unidirectional block formation when the electrode is located near a scarred region [23]. Moreover, some computational studies have recently focused on the prediction of arrhythmias in several scenarios, such as acute ischemia [24, 25, 26, 27], chronic ischemia [28, 29] and non-ischemic fibrosis [30].

In this study, we present an original computational study where a personalized CRT model is applied to patients with non-ischemic fibrosis to assess the validity of the computational prediction of Ventricular Tachycardia (VT) during BiV-CRT. To the best of our knowledge, this is the first attempt to successfully predict VT in BiV-CRT patients by means of a computational tool.

To this aim, for each patient we personalize the electrical properties of the computational model using activation times coming from an Electro-Anatomical Mappings System (EAMS) [31, 32], and we propose an ionic current model for non-ischemic myocardial fibrosis regions. Then, we run electrophysiology simulations employing a 𝕊1 − 𝕊2 stimulation protocol adapted to BiV-CRT cases, to assess the propensity of VT formation of the patient. To validate our outcomes, we compare the predicted VT risk with follow-up at disposal for the patient.

## 2 Materials and methods

### 2.1 Clinical data

Our study includes three CRT patients, P6, P10, P11, numbered as in [33], that were treated at Santa Maria del Carmine Hospital, Rovereto (TN), Italy. These patients suffered from ventricular dyssynchrony caused by Left Bundle Branch Block (LBBB). The etiology of the LV dysfunction was non-ischemic in all the three patients. In addition, all feature a fibrotic region detected by imaging.

For each patient, the available clinical data comprise:

- *The device characteristics and the properties of stimulation*. The patients have undergone the implantation of a BiV-CRT consisting of three electrodes: the first one positioned in the right atrium, the second one at the right ventricle apical endocardium, and the last one at the LV epicardium via the coronary sinus. Specifically, the LV lead was targeted to the *Latest Electrically Activated Segment* (LEAS) [34]. For each patient, a specific VV-delay between the electrodes was set, with the right lead delayed with respect to the left one (the values of the delays are reported in Table 1).
- *Magnetic Resonance Imaging (MRI) acquisitions*. These data consist in Cine Steady-State Free Precession (Cine-SSFP) short-axis sequences and in Phase Sensitive Inversion Recovery (PSIR) sequences with Late Gadolinium Enhancement (LGE), which is a key factor in assessing both ischemic and non-ischemic cardiomyopathies.
- *Electro-Anatomical Mappings System (EAMS) data*. These provide the electrical activation times obtained during sinus rhythm (pre-operative scenario) on all mapped points performed during CRT procedure. Notice that EAMS acquisition is obtained by cardiologists during the CRT procedure and no additional invasive mapping is required for our purposes.
- *Follow-up data*. Continuous monitoring via the BiV-CRT device provides detailed registration and documentation of arrhythmic events. The significant clinical follow-up for the three patients is reported in Table 1. Based on these documented events, for the purpose of this study patients P6 and P10 are classified as arrhythmic, whereas patient P11 as non-arrhythmic.

**Table 1:** Values of delay adjusted by clinicians (VV-delay) and clinical follow-up for each patient. Acronyms: Ventricular Tachycardia (VT); Antitachycardia Pacing (ATP).

| Patient | VV-delay [ms] | Follow-up |
| --- | --- | --- |
| P6 | 30 | Unsustained VT (6 seconds) |
| P10 | 20 | VT terminated with ATP and shock |
| P11 | 0 | No events |

### 2.2 Reconstruction of patient-specific geometries

To define the computational domain for the electrophysiology problem, the initial step involves extracting the LV geometries and the myocardial fibrotic regions from the patient’s MRI data. The LV geometry is extracted from Cine-SSFP images thanks to their high resolution, while the fibrosis geometry is extracted from LGE-MRI, which clearly delineates the scar shape. To ensure precise anatomical alignment of the functional (Cine-SSFP) and fibrosis (LGE-MRI) planes, images are synchronized at mid-diastole, which is the conventional acquisition phase for LGE-MRI images.

More specifically, starting from the MRI imaging, the segmentation of the ventricles is performed using the free, open-source software *3D Slicer* [35]. Subsequently, *VMTK* (Vascular Modeling Toolkit) [36, 37] is employed to incorporate the fibrotic tissue into the left ventricle (LV) geometry, completing the 3D reconstruction. Final pre-processing is achieved using *Paraview* [38], an open-source data analysis and visualization application, which allows for the spatial identification and differentiation of the two regions: the healthy tissue and the fibrotic tissue. The resulting reconstructed geometries are shown in Figure 1, right. In the same figure, on the left, we report a detailed bullseye representation, where for each of the 17 segments we identify the endocardial, the myocardial and the epicardial region, each coloured in grey in presence of fibrosis. This allows us to have more specific information about the presence of the fibrosis across tha transmural depth.

**Figure 1:**
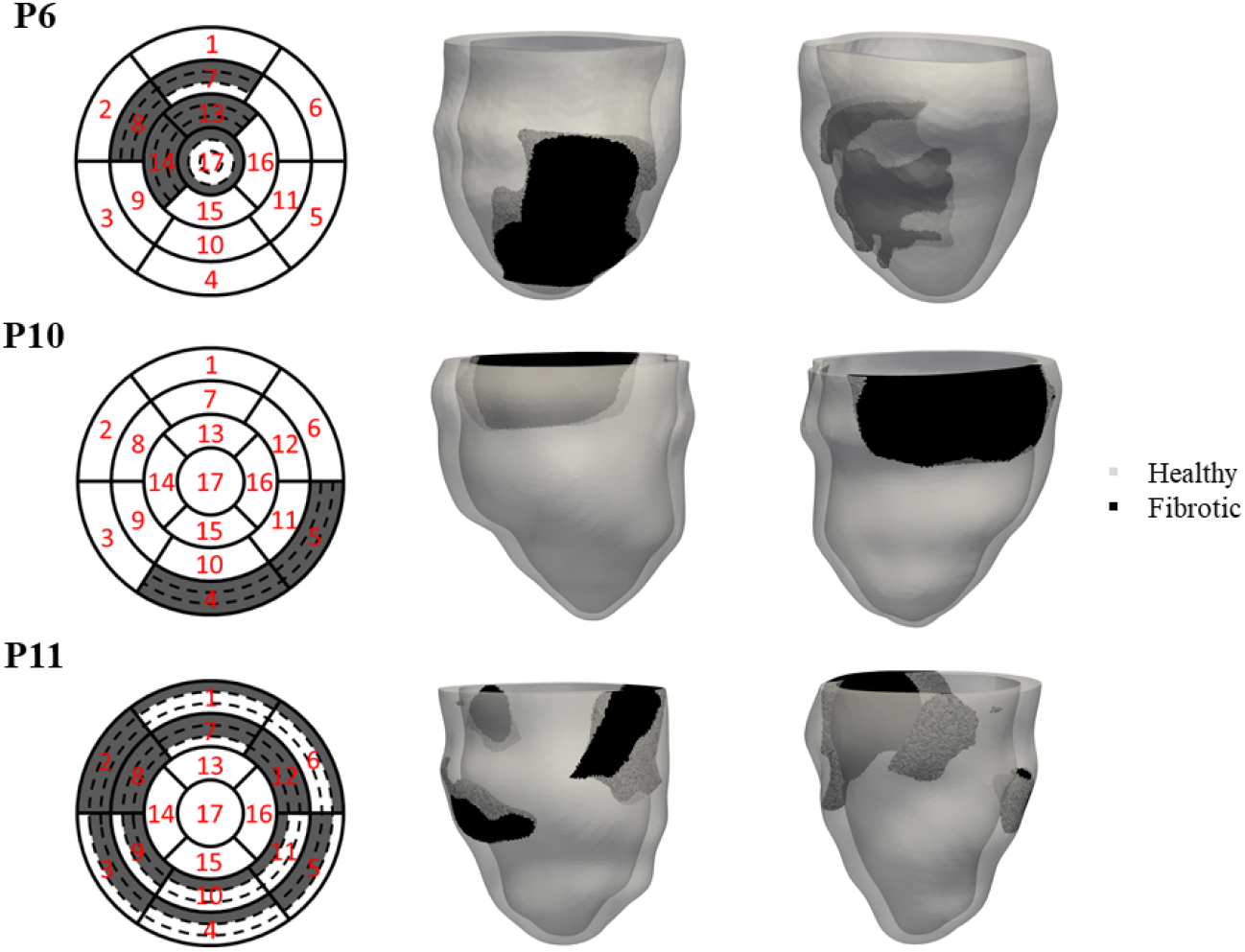
3D Reconstruction of the LV and the fibrosis for patients P6, P10, and P11. The fibrotic regions are derived from the clinicians’ bullseye plots, which are adapted to represent the three distinct layers (endocardial, myocardial and epicardial) within the affected segments.

Using VMTK, we generate for each patient the finite-element mesh consisting of tetrahedral elements. The average element dimension is set to *h* = 0.7*mm* which using in combination with P2 Finite Elements (see Section 2.3) guarantees the required resolution for accurate electrophysiology simulations. The resulting mesh for a representative patient is shown in Figure 2.

**Figure 2:**
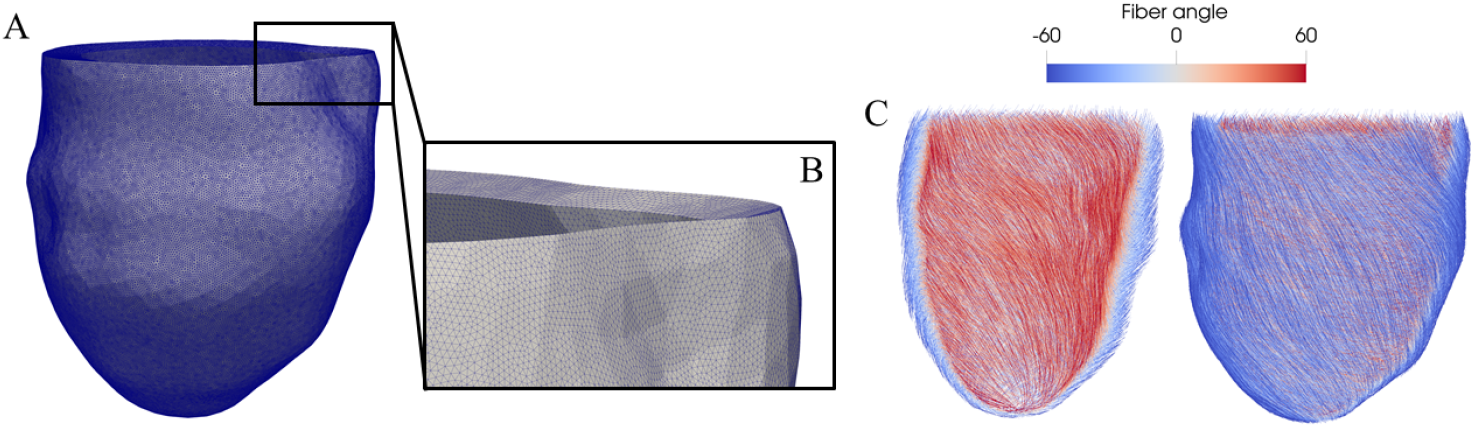
Finite-elements mesh and myocardial fiber orientation for patient P6. (A) The complete 3D reconstructed geometry, discretized using tetrahedral elements. (B) A zoomed-in view highlighting the resulting tetrahedral mesh structure. (C) Visualization of the computed myocardial fiber orientation, where the color bar indicates the fiber angle in degrees with respect to the circumferential direction.

Finally, the complex fibers-sheets architecture that characterizes the myocardium is built using a Laplace-Dirichlet rule-based numerical method [39, 40] (see Figure 2 for a representative patient).

As regards the computational setup for the clinical BiV-CRT protocol, we follow the methodology described in [21]. Specifically, we neglect the atria and thus we consider only the left and right leads. The former one is located at LEAS, whereas the latter on the LV epicardium. The specified lead locations are visible in Figure 3.

**Figure 3:**
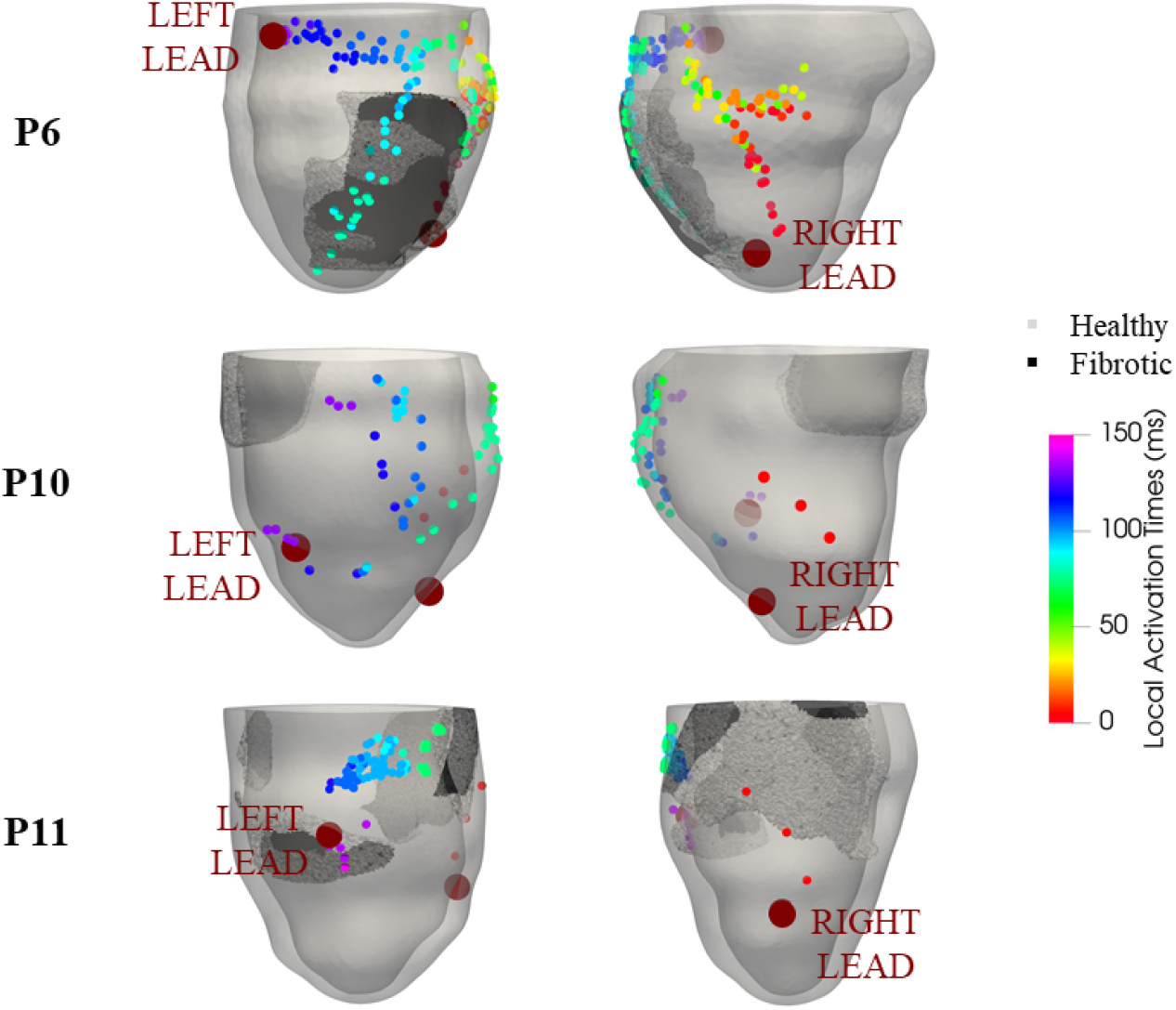
Pre-operative Local Activation Time (LAT) from EAMS and pacing sites for the three patients. The background geometries show fibrotic and healthy tissue, while the colored dots show the pre-operative LAT distribution. Large burgundy spheres mark the pacing electrode locations: left column shows the left lead placed at LEAS; right column shows the right lead applied at the interventricular septum.

### 2.3 Overview of the computational model

We consider the personalized computational model proposed for BiV-CRT patients in [21], applied here to the data and computational meshes of the new cases described in the previous sections. This model has been validated in [21] allowing to well predict electrophysiology patterns not used in the personalization procedure.

The model is based on the monodomain equation for the transmembrane potential [41, 42] combined with the Ten Tusscher-Panfilov (TTP06) model for ionic currents [43], adapted to the fibrotic case as detailed in Section 2.4. Starting from the activation times obtained from EAMS and accordingly to the calibration procedure introduced in [31, 32], we personalize the values of the monodomain conductivities for each patient, differentiating their values in the healthy and fibrotic regions, see Table 2.

**Table 2:** Values of the personalized conductivity along the fibres (*σ*_*f*_), sheets (*σ*_*s*_), and normal direction (*σ*_*n*_), normalized with respect to the surface-to-volume ratio and the transmembrane capacitance. The values are in [10^−4^*m*^2^*/ms*]. Subscript H stands for *healthy region*; subscript F stands for *fibrotic region*.

| Patient | $\sigma_{f,H}$ | $\sigma_{f,F}$ | $\sigma_{s,H}$ | $\sigma_{s,F}$ | $\sigma_{n,H}$ | $\sigma_{n,F}$ |
| --- | --- | --- | --- | --- | --- | --- |
| P6 | 1.75 | 1.40 | 0.28 | 0.24 | 0.07 | 0.05 |
| P10 | 1.42 | 1.02 | 0.35 | 0.25 | 0.08 | 0.06 |
| P11 | 1.42 | 0.60 | 0.37 | 0.15 | 0.08 | 0.03 |

All the numerical experiments are run using `life`^X^ [44], an open-source C++ library for the simulation of the heart functions, developed at MOX, Dipartimento di Matematica, in collaboration with LaBS, Dipartimento di Chimica, Materiali e Ingegneria Chimica “Giulio Natta” (Politecnico di Milano). See [45] for details on electrophysiological solvers and the relative open-source binary tool. We use a time discretization parameter equal to Δ*t* = 5 · 10^−5^ *s* [46] and ℙ^2^ Finite-Elements for space discretization, for which the average value of *h* = 0.7*mm* in the meshes reported in the previous section guarantees a sufficient accuracy in approximating the transmembrane potential dynamics [27].

### 2.4 Ionic current model for non-ischemic fibrosis

In this section, we present the mathematical model for the ionic currents adapted to the case of a fibrotic non-ischemic region. To this aim, we refer to [47] where the authors proposed a fibroblast cell activation model suitably adapting the standard physiological TTP06 model [43]. Instead of including the fibroblast cell activation model, here we prefer to suitably change the parameters of the TTP06 model in the fibrotic region in order to reproduce the specific morphology of the fibrotic action potential (AP) reported in [47], see Figure 4. This could be interpreted as an average behaviour among the different fibrotic cells of the region. Notice, as expected, a reduction of the voltage spike, an increment of the resting potential, and a shortening of AP duration. Specifically, these changes amounted to approximately 27.6%, 21.0%, and 19.8%, respectively.

**Figure 4:**
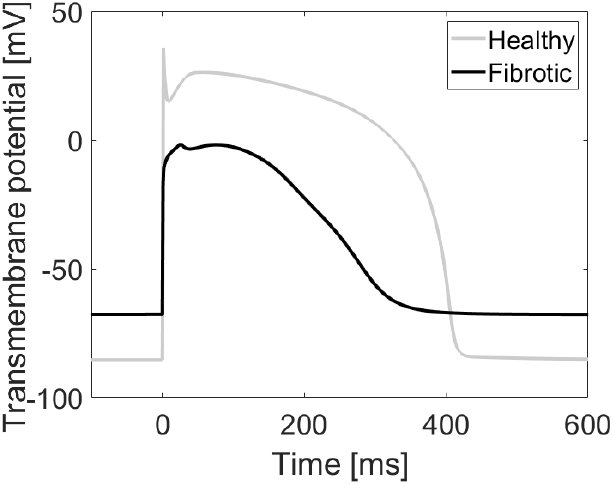
Action potential morphologies for the healthy (grey) and fibrotic (black) regions. These morphologies are obtained using our adapted TTP06 model, which simulates the average electrophysiological behavior of each respective region.

In Table 3 we report the values of the adapted parameters in our TTP06 model.

**Table 3:** Values of the parameters used in the adapted TTP06 model. For the other parameters, not reported here, we use for the fibrotic region the same values of the healthy one, as in [43]. [*K*^+^]_*o*_: Resting concentration of external potassium ion [*mmol/L*]; *G*_*kr*_: Maximal conductance related to potassium current *I*_*Kr*_ [*nS/pF*]; *G*_*to*_: Maximal conductance related to potassium current *I*_*to*_ [*nS/pF*]; *G*_*K*1_: Maximal conductance related to potassium current *I*_*K*1_ [*nS/pF*]; *G*_*pK*_: Maximal conductance related to potassium current *I*_*pK*_ [*nS/pF*]; *G*_*Na*_: Maximal conductance related to sodium current *I*_*Na*_ [*nS/pF*]; *G*_*CaL*_: Maximal conductance related to calcium current *I*_*CaL*_ [*nS/pF*]; *k*_*NaCa*_: Maximal scaling coefficient related to sodium-calcium exchanger current [*pA/pF*].

| Region | $[K^+]_o$ | $G_{Kr}$ | $G_{to}$ | $G_{K1}$ | $G_{pK}$ | $G_{Na}$ | $G_{CaL}$ | $k_{naca}$ |
| --- | --- | --- | --- | --- | --- | --- | --- | --- |
| Healthy | 5.4 | 0.153 | 0.3 | 5.4 | 0.15 | 14.8 | 0.000040 | 1000 |
| Fibrotic | 10.0 | 0.015 | 8.0 | 1.0 | 0.05 | 25.0 | 0.000006 | 500 |

### 2.5 Virtual stimulation procedure

In this work, the stimulation procedure is adapted from established virtual stimulation protocols used in computational studies of VT formation [26, 27] to specifically model the BiV-CRT case.

The protocol for each simulation consists of two sequential steps:

- *S1: CRT pacing*. The simulation is initiated by applying two electrical impulses corresponding to the left and right ventricular leads. The first impulse is delivered at time *t* = 0*ms*, while the second impulse is applied after a time delay specified in Table 1. The precise locations of these virtual leads are illustrated in Figure 3.
- *S2: Ectopic impulse*. Following the CRT pacing sequence, a single ectopic impulse is applied in the vicinity of the fibrotic region. This action is designed to simulate a pathological trigger arising from cells that exhibit abnormal automaticity or activity triggered by the presence of an anatomical obstacle, such as myocardial fibrosis [48].

To assess the arrhythmogenic risk of each patient, we execute multiple simulations for a set of ten distinct ectopic sites selected around the fibrotic region. The aim of this study is to quantify a patient’s vulnerability to arrhythmias by considering the number of ectopic impulses that successfully generate at least one reentry loop. Indeed, the formation of a reentry loop is considered a strong computational signal indicating the possible initiation of reentrant VT.

## 3 Results

The results of the patient-specific electrophysiological simulations are presented here, structured in two sections: first, we analyze the resulting electrical activation pattern of the left ventricle under standard BiV-CRT pacing (Section 3.1); second, we detail the arrhythmogenic risk assessment derived from the virtual ectopic stimulation protocol (Section 3.2).

### 3.1 Activation pattern under BiV-CRT pacing

Numerical simulations of the personalized models are run under the BiV-CRT virtual pacing protocol (Section 2.5). In this first analysis, we simulate the post-operative scenarios without ectopic beats to assess the resulting electrical pattern under therapeutic pacing. These simulations incorporate all patient-specific model components, including the personalized monodomain conductivities (Table 2) and the adapted TTP06 model used to characterize the electrical behavior of the fibrotic tissue (Section 2.4). Figure 5 shows the propagation of the electrical impulses activated by the BiV-CRT with respective activation and repolarization maps.

**Figure 5:**
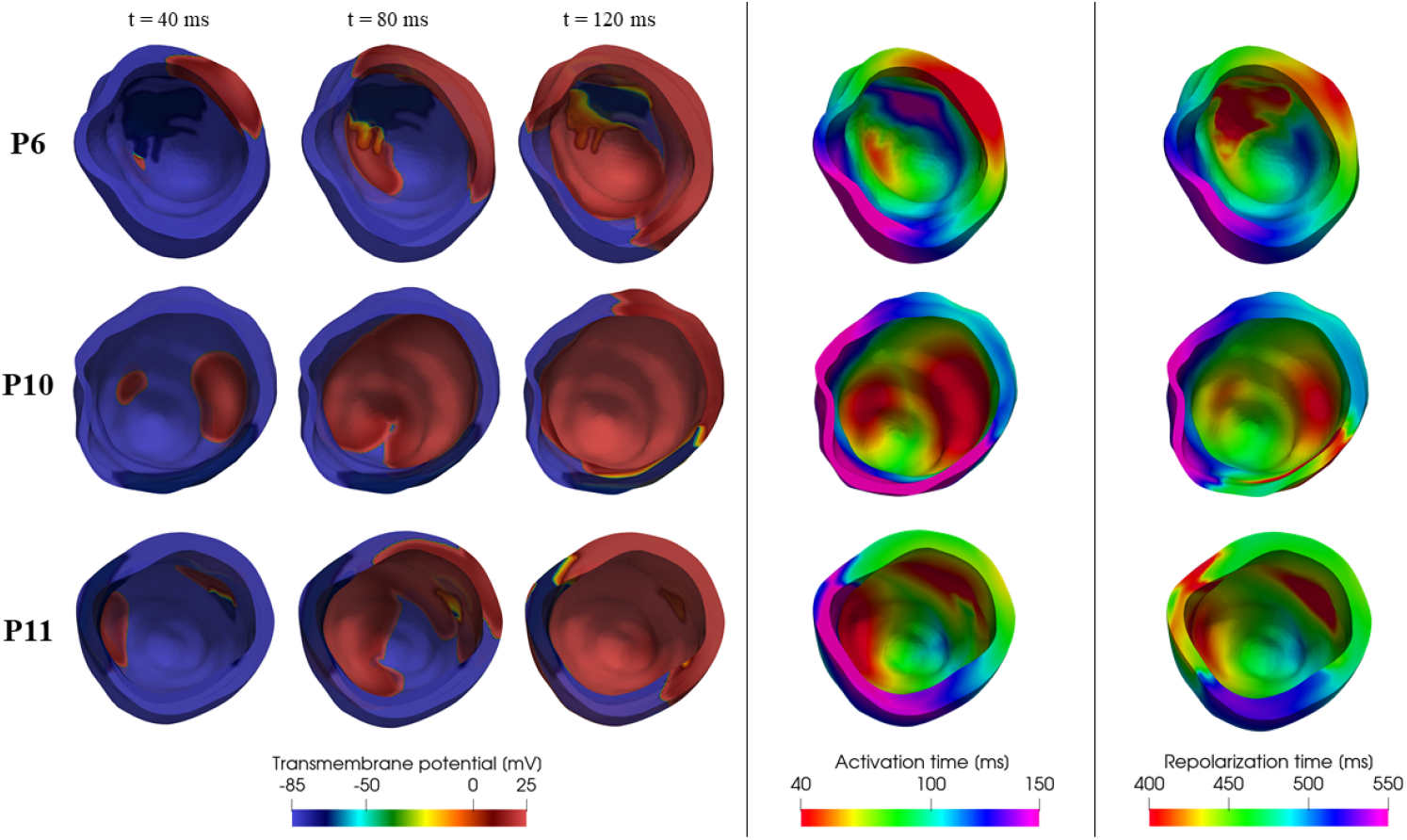
Electrophysiological maps for patients P6, P10, and P11 (rows) under BiV-CRT pacing. Left columns: Transmembrane potential propagation at *t* = 40, 80, and 120 ms. Center column: Activation time maps. Right column: Repolarization time maps.

We notice that our simulations are able to reproduce the double pacing at the apex and at the epicardial veins characteristic of BiV-CRT. Even if not directly validated against BiV-CRT EAMS measurements (not available), as discussed in Section 2.3, our computational model were validated in [21] against EAMS measurements in the case of right pacing; thus, we can assume that also in the case of biventricular pacing, our model features a high level of accuracy and prediction.

### 3.2 Arrhythmogenic risk assessment

The personalized models are then subjected to the virtual stimulation protocol described in Section 2.5, where a total of 10 distinct ectopic impulses are applied around the fibrotic region for each patient, see Figure 6 (orange bullets). In the same figure we depict in red those ectopic impulses that generate a reentry loop. The results regarding the arrhythmogenic risk of each patient together with the clinical follow-up indication are summarized in Table 4.

**Table 4:** Results of the arrhythmogenic risk assessment. The second column reports the total number of distinct ectopic sites (out of ten) that resulted in the formation of a reentry loop. The third column reports our model’s prediction based on the reentry count compared to the documented clinical status (last column).

| Patient | Arrhythmogenic ectopic sites / 10 | Clinical follow-up |
| --- | --- | --- |
| P6 | 6 | Arrhythmic |
| P10 | 6 | Arrhythmic |
| P11 | 3 | Non-arrhythmic |

**Figure 6:**
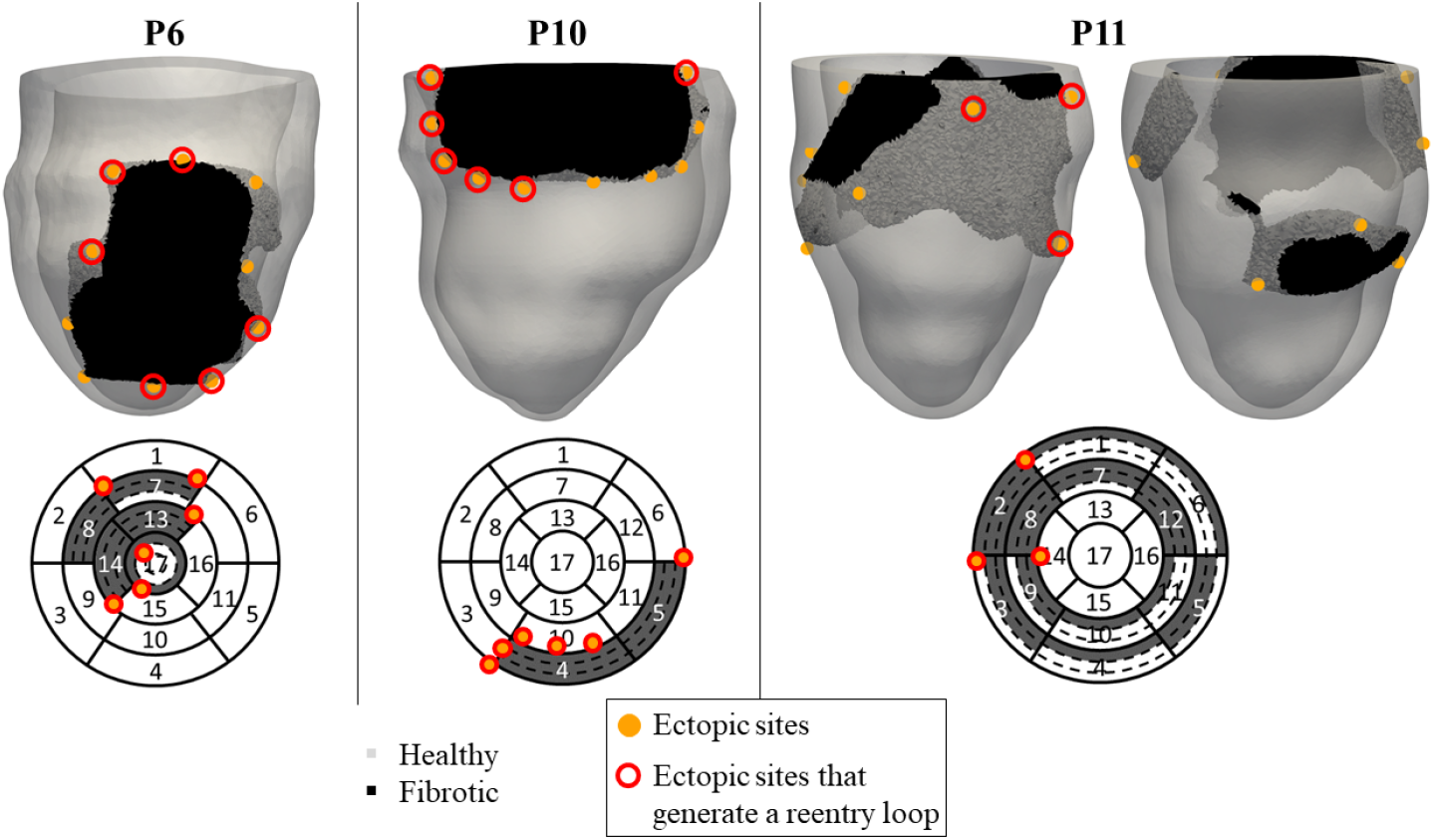
Localization of the 10 ectopic sites across three patients (P6, P10, P11). The 3D ventricular geometries illustrate all ten ectopic sites (orange circles). Sites that generate a reentry loop are highlighted with a red ring. The corresponding bullseye plots show only the ectopic sites that are arrhythmogenic, indicating their segmental position within the ventricle.

The computational study shows that the number of ectopic impulses that generate a reentry loop is higher for patients P6 and P10 (6 and 6 out of 10, respectively) compared to patient P11 (3 out of 10). This observed pattern is consistent with the documented clinical status of patients P6 and P10 as arrhythmic, and patient P11 as non-arrhythmic (Table 1 and 4). Figure 7 provides a visual example of a successful reentry loop initiated by an ectopic stimulus in P6 and P10, with also an example of a decaying ectopic impulse in P11.

**Figure 7:**
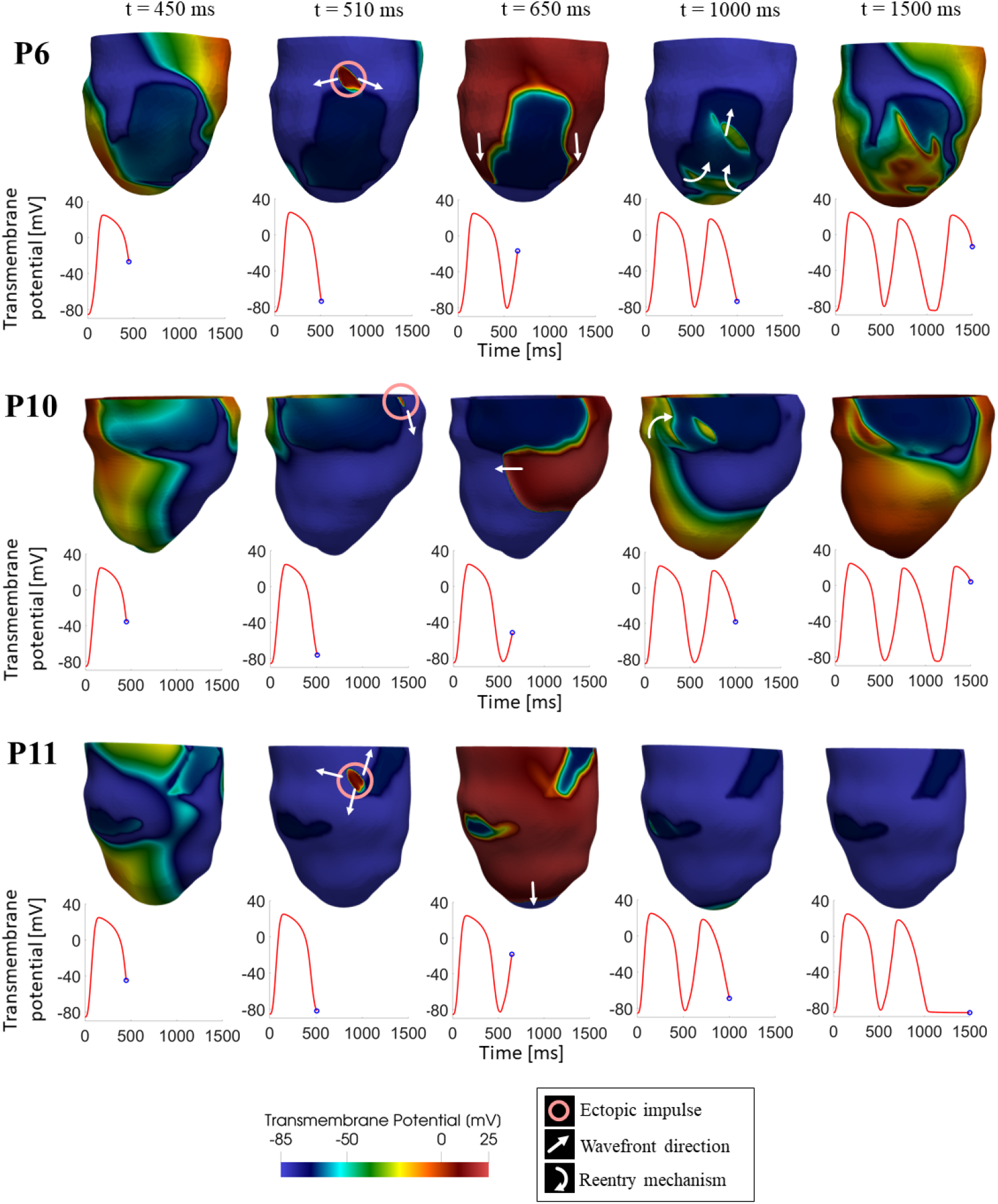
Epicardial transmembrane potential propagation at five different time istants, during BiV-CRT in presence of an ectopic impulse originating at the repolarization tail. In P6 and P10, this ectopic impulse successfully initiates a sustained reentry loop, with the reentrant activation visible as the third spike in the potential trend plots. In P11, the ectopic impulse decays and does not generate reentries.

## 4 Discussion

The present study demonstrated a novel application of a patient-specific computational cardiac model to assess the risk of VT in patients undergoing BiV-CRT who also suffer from non-ischemic myocardial fibrosis.

The personalized computational setup, which incorporated patient-specific monodomain conductivities (Table 2) and an adapted Ten Tusscher-Panfilov (TTP06) ionic current model for non-ischemic fibrosis (Section 2.4), reliably reflected the individual conduction properties affected by the pathology. This approach was essential for capturing the altered electrical behavior within the arrhythmogenic substrate (Figure 4).

It is well-established that myocardial fibrosis is a complex, heterogeneous substrate that creates surviving myocardial bundles, which may be primary drivers of reentrant arrhythmias [49, 50]. While some computational studies incorporated these conduction channels by manually designing them [51, 52], we are not aware of the inclusion in this type of studies of a patient-specific design of the channels in the fibrosis. Accordingly, in computational studies where patient-specific reconstruction of fibrosis were considered, they were all accompanied by the modeling assumption of absence of conduction in such a region [15, 19]. Since our primary goal was to develop a predictive tool relying exclusively on data acquired during the standard clinical CRT procedure (LGE-MRI and, possibly, EAMS) and since specific, non-standard acquisitions would be needed to detect patient-specific fibrotic channels (such as Ultra High-Density mapping), we tried to overcome the limitation introduced by other computational studies by using the action potential reported in Figure 4 for all the cells of the fibrosis. This AP should then be thought as an “average” value among the fibrotic cells, which, if on the one hand did not allow us to identify the channels, at least on the other hand better characterized the electrophysiology in fibrosis compared to considering all its cells as non-conducting.

The BiV-CRT simulations in absence of any ectopic beat allowed us to characterize the typical electrical propagation patterns under this pacing conditions. These results confirmed the expected double activation wave initiated at the apex and the epicardial veins, demonstrating the model’s capability to accurately simulate the post-operative electrical state under pacing conditions (Figure 5). Specifically, the activation maps reveal heterogeneous conduction among the three patients, consistent with their personalized structural and electrical properties. Furthermore, the repolarization maps highlight significant differences in the distribution and dispersion of the repolarization time, indicating patient-specific electrophysiological vulnerabilities even under therapeutic pacing conditions.

The primary finding of this study was the predictive coherence between the computationally assessed VT risk obtained introducing ectopic beats during the BiV-CRT pacing and the long-term clinical follow-up of BiV-CRT patients with non-ischemic fibrosis (Table 4). Patients P6 and P10, who were clinically classified as arrhythmic, showed significantly higher arrhythmogenic propensity, with 6 ectopic impulses out of 10 that form a reentry loop, compared to the non-arrhytmic patient P11 (only 3 ectopic sites with loop formations).

It is interesting to notice the figure-of-eight morphology of the reentrant loops (Figure 7). This structure is typically observed in Ventricular Tachycardia (VT) events, which are characterized by organized circuits often linked to the presence of an anatomical obstacle like scars [53]. By successfully simulating VT, our model showed clear coherence with the clinical follow-up of the studied patients, in which chaotic events such as Ventricular Fibrillation (VF) were not observed. The initiation of these organized loops consistently occurred around the fibrotic region, confirming that changes in conductivity and altered cellular properties modified the substrate to induce a high refractory period and subsequent organized reentry.

Although the number of cases is very limited, we try here to provide a possible rationale behind the persistent formation of reentries. From the bullseye representation of Figure 1, we notice that P6 and P10 (those who experienced arrhytmogenic episodes) are characterized by a fibrosis distribution which is confined in a specific region. This probably allowed the signal to propagate across a wide region and, due to the refractoriness heterogeneities, to form loops. On the other side, for P11 (who did not experience arrhythmogenic episodes) the fibrotic region is distrubuted along a large region and at the same time its volume is larger than P6 and P10. This heterogeneity might not have created the substrate for a reentry loop, due to the absence of sufficiently long pathways. Notice that these findings (arrhythmogenesis risk related to the substrate heterogeneity rather than to its volume) are in agreement with the observations provided in [27] for the case of acute ischemia.

Another observation in this direction is given by noticing that all the ectopic beats leading to a reentry (see Figure 6, bottom) are located, for all the three patients, adjacent to segments with full transmural fibrosis (apart from segment 7 in P6).

To conclude, in this study, we have showcased a practical application of a methodology to predict VT formation under BiV-CRT pacing. Our intention was not to conduct extensive statistical analyses, given the relatively modest sample size. Instead, our focus was on presenting a proof-of-concept to highlight the potential of the methodology. The proposed pipeline seems particularly well-suited for systematic application to patient-specific cases, since it requires only standard medical data acquisitions. This methodology could serve as the foundation for a future routine procedure aimed at assessing the arrhythmogeic risk during BiV-CRT pacing, in order to provide useful information to cardiologists on possible treatments (for example farmacological or CRT-D).

### 4.1 Limitations

Despite these encouraging results, the study has some limitations:

- *Small patient cohort*. One of the limitations of the study is the minimal patient cohort (*n* = 3). While the high predictive consistency is noted, it is statistically insufficient to establish clinical robustness. Rigorous validation on a significantly larger, multi-center dataset is necessary.
- *Lack of intramural and apex-base heterogeneity*. The TTP06 ionic model is applied uniformly across the ventricular wall thickness. This neglects physiological variations (e.g., endocardial, myocardial and epicardial properties) and the apex-to-base gradient in repolarization. Ignoring this dispersion of repolarization may lead to an inaccurate characterization of the patient-specific vulnerability window, especially under non-physiological BiV-CRT pacing.
- *Stimulation protocol*. The arrhythmogenic assessment relied on the analysis of only one ectopic beat as trigger of VT formation (𝕊1 − 𝕊2 stimulation). Future work should explore the case of a possible train of ectopic beats that originate in a location to comprehensively map the patient vulnerability.
- *Lack of BiV-CRT EAMS validation*. While the model was personalized against pre-operative EAMS (sinus rhythm) and validated in right pacing conditions (see [21]), the resulting post-operative BiV-CRT activation patterns were not directly validated against EAMS data acquired under biventricular pacing conditions, since these data were not available. Future analyses could focus on filling this gap provided that EAMS will be performed during BiV-CRT.
- *Absence of the His-Purkinje System (HPS)*. Since the origin of the signal is provided in our cases by the BiV-CRT device, we did not include the Purkinje system in our computational model, see e.g. [54, 55].

However, in BiV-CRT pacing, the signal could enter antidromically the Purkinje network and this could have an impact on the loops’ formation [56, 57]. For this reason, future studies could include a Purkinje system modeling to assess its influence on VT development during BiV-CRT stimulation.

## 5 Translational perspective

Computational methods have been widely used in recent years in order to study and optimize the CRT procedure, particularly regarding the challenging topic of optimal electrode positioning and VV-delay determination. While the research community has made considerable improvements in this direction, it is still far from providing a device implemented in the clinical routine, that could assist clinicians in their decision-making process. With this work, we contributed in this direction by introducing a multi-objective criterion for personalized CRT. Our framework highlighted the potential of computational electrophysiology as a clinically relevant tool for risk stratification, non-invasively identifying patients with increased vulnerability to ventricular tachycardia during BiV-CRT pacing. Moreover, this model was complementary to existing optimization strategies, allowing clinicians to simultaneously consider pacing criteria and recognize cases with arrhythmic risk. Ultimately, this approach could pave the way for routine computational assessment of the arrhythmic substrate, improving the safety and therapeutic efficacy of patients undergoing CRT.

## Data Availability

All data produced in the present study are available upon reasonable request to the authors

## 6 Acknowledgments

AC, SP, CV are members of the INdAM group GNCS “Gruppo Nazionale per il Calcolo Scientifico” (National Group for Scientific Computing).

CV has been partially supported by: i) the European Union-Next Generation EU, Mission 4, Component 1, CUP: D53D23018770001, under the research project MIUR PRIN22-PNRR n.P20223KSS2, “Machine learning for fluid structure interaction in cardiovascular problems: efficient solutions, model reduction, inverse problems”, ii) the Italian Ministry of Health within the PNC PROGETTO HUB LIFE SCIENCE - DIAGNOSTICA AVANZATA (HLS-DA) “INNOVA”, PNCE3-2022-23683266–CUP: D43C22004930001, within the “Piano Nazionale Complementare Ecosistema Innovativo della Salute” - Codice univoco investimento: PNCE3-2022-23683266; iii) the Italian research project MIUR PRIN22 n.2022L3JC5T “Predicting the outcome of endovascular repair for thoracic aortic aneurysms: analysis of fluid dynamic modeling in different anatomical settings and clinical validation”; iv) Italian Ministry of Health within the project “CAL.HUB.RIA” - CALABRIA HUB PER RICERCA INNOVATIVA ED AVANZATA. Code: T4-AN-09, CUP: F63C22000530001.

SP is partially supported by INNOVA and acknowledges the support of the MUR, Italian Ministry of University and Research, grant Dipartimento di Eccellenza 2023–2027. SP acknowledges the INdAM GNCS project CUP E53C24001950001.

AC is funded by INNOVA.

The authors acknowledge the CINECA award under the ISCRA initiative, for the availability of high-performance computing resources and support.

## 7 Declarations

### Ethical approval

Ethical Review Board approval was obtained (R250/15-CCM 262). All procedures performed in studies involving human participants were in accordance with the ethical standards of the institutional and/or national research committee and with the 1964 Helsinki declaration and its later amendments or comparable ethical standards.

### Informed consent

Written informed consent was obtained from all subjects (patients) in this study.

